# Real-World Outcomes of Virtual Medical Nutrition Therapy in Dyslipidemia, Hypertension, or Diabetes

**DOI:** 10.64898/2026.10.01.26364516

**Authors:** Rebecca N. Adams, Chloe Bollinger, Dariush Mozaffarian

**Affiliations:** Clinical Research, Nourish, Inc, New York, NY; Food Is Medicine Institute, Friedman School of Nutrition Science and Policy, Tufts University, Boston, Massachusetts; Division of Cardiology, Tufts Medical Center, and Tufts University School of Medicine, Boston, Massachusetts

## Abstract

**Importance:** Dyslipidemia, hypertension, and diabetes are highly prevalent conditions for which medical nutrition therapy (MNT) is guideline-recommended, yet access remains limited, and real-world evidence on technology-enabled MNT models at scale is sparse.

**Objective:** To characterize lipid, blood pressure, and glycemic outcomes among adults receiving virtual MNT (vMNT) across three cardiometabolic conditions.

**Design:** Retrospective cohort study of adults with a first Nourish appointment before November 2025, followed for 6 months.

**Setting:** National vMNT platform.

**Participants:** Adults meeting enrollment criteria for dyslipidemia, hypertension, or diabetes and remaining engaged in care with ≥1 follow-up measurement at 3 or 6 months. Final cohorts: dyslipidemia (n=6,501), hypertension (n=13,704), diabetes (n=2,444); 18,965 unique patients overall.

**Exposure:** vMNT delivered by a registered dietitian via telehealth visits, with technology-enabled support between visits.

**Main Outcomes and Measures:** Outcomes were LDL cholesterol, total cholesterol, HDL cholesterol, and triglycerides (dyslipidemia); systolic and diastolic blood pressure (SBP, DBP) (hypertension); and HbA1c (diabetes), assessed at enrollment, 3, and 6 months. Weight was a secondary outcome. The proportion achieving guideline-recommended targets (HbA1c <7.0%, SBP <130 mmHg) was also assessed.

**Results:** Among 18,965 patients (mean [SD] age, 42.9 [11.6] years; 15,770 [83.2%] female), significant changes from baseline to 6 months included: LDL cholesterol (−14.0 mg/dL [SE, 0.56]); total cholesterol (−19.4 mg/dL [SE, 0.61]); HDL cholesterol (−0.5 mg/dL [SE, 0.15]); triglycerides (−28.6 mg/dL [SE, 1.06]); SBP (−5.5 mmHg [SE, 0.16]); DBP (−4.5 mmHg [SE, 0.11]); and HbA1c (−1.37 percentage points [SE, 0.04]) (all P<.001). Among those with elevated SBP, 57.3% achieved <130 mmHg by 6 months; among those with elevated HbA1c, 54.7% achieved <7.0%. Outcomes among patients with stable medication regimens were similar to the full cohort.

**Conclusions and Relevance:** In this study of 18,965 patients, vMNT delivered through a technology-enabled care model was associated with clinically meaningful improvement across three prevalent cardiometabolic conditions. These findings support technology-enabled virtual care as a scalable approach to delivering effective MNT.

---

Dyslipidemia, hypertension, and diabetes are core components of cardiovascular-kidney-metabolic (CKM) syndrome^1^ and highly prevalent among US adults, producing significant cardiovascular morbidity, mortality, and healthcare spending.^2^ Nutritional intervention is a cornerstone of their management, including guideline-recommended medical nutrition therapy (MNT) provided by a registered dietitian (RD) which effectively improves these risks in controlled trials.^3–6^ However, access to MNT often remains limited in traditional care settings, reflecting inadequate and geographically uneven RD staffing, low provider referral rates, and inconsistent payer reimbursement.^6–8^

Telehealth-delivered MNT offers one way to expand access beyond traditional in-person care by reducing geographic and logistical barriers.^9^ Telehealth-delivered MNT has been evaluated in a growing number of small randomized trials, generally demonstrating meaningful improvements across hypertension, diabetes, and broader cardiovascular risk factors, with effect sizes similar to those reported in in-person MNT studies.^10–12^ Building on this foundation, one emerging model pairs an RD as the primary clinician with a broader technology platform that extends support between visits — allowing patients to message their RD directly, access educational resources, and receive AI-generated feedback in response to logged dietary and health data, extending beyond the single-encounter structure of a typical telehealth or in-person appointment. However, prior real-world evidence from this type of model remains limited, generally focused on weight loss rather than condition-specific outcomes.^13^ Additional investigation of technology-enabled virtual MNT’s association with these condition-specific cardiometabolic outcomes is needed to understand the potential role of this scalable strategy in improving cardiometabolic health.

To address this gap, we conducted a retrospective, real-world study of patients receiving usual-care virtual MNT (vMNT) through Nourish, a platform delivering this type of technology-enabled care. Our outcomes were lipid, blood pressure, and glycemic levels at 3 months and 6 months, including the proportion of patients achieving guideline-recommended clinical targets. We additionally examined outcomes among patients with stable medication regimens to characterize the contribution of vMNT independent of concurrent pharmacotherapy, informing the potential of this scalable care model to expand effective MNT delivery.

## Methods

### Study Design and Population

This retrospective observational cohort study used data from the Nourish clinical registry, a de-identified dataset derived from the electronic health records (EHRs) of adults receiving care through Nourish. Patients accessed Nourish through multiple pathways, including provider and health system referrals, direct marketing, and national health plan coverage. The study was determined exempt by the Biomedical Research Alliance of New York (BRANY) institutional review board.

The study comprised three condition-specific cohorts, with primary outcomes of LDL cholesterol (LDL-C), total cholesterol, HDL cholesterol (HDL-C), and triglycerides among adults with dyslipidemia; systolic and diastolic blood pressure (SBP, DBP) among adults with hypertension; and HbA1c among adults with diabetes. Weight was a secondary outcome across all three cohorts. Outcomes were assessed at enrollment, 3, and 6 months, with 6 months as the primary timepoint.

Eligible patients were adults aged 18 years or older with a first Nourish appointment before November 2025 (eFigure 1). Following exclusion of physiologically implausible values from clinical outcome data, three condition-specific cohorts were defined based on clinical values at enrollment: dyslipidemia (LDL cholesterol ≥130, total cholesterol ≥200, or triglycerides ≥150 mg/dL); hypertension (SBP ≥130 or DBP ≥80 mmHg); and diabetes (HbA1c ≥6.5%). Cohorts were not mutually exclusive; patients meeting criteria for more than one condition were included in each applicable cohort. We restricted analyses to patients who were engaged with their RD (a recorded visit ≥150 days after their first appointment) with ≥1 follow-up outcome measurement at 3 or 6 months, yielding 18,965 unique patients: 6,501 with dyslipidemia, 13,704 with hypertension, and 2,444 with diabetes.

**Figure 1.**
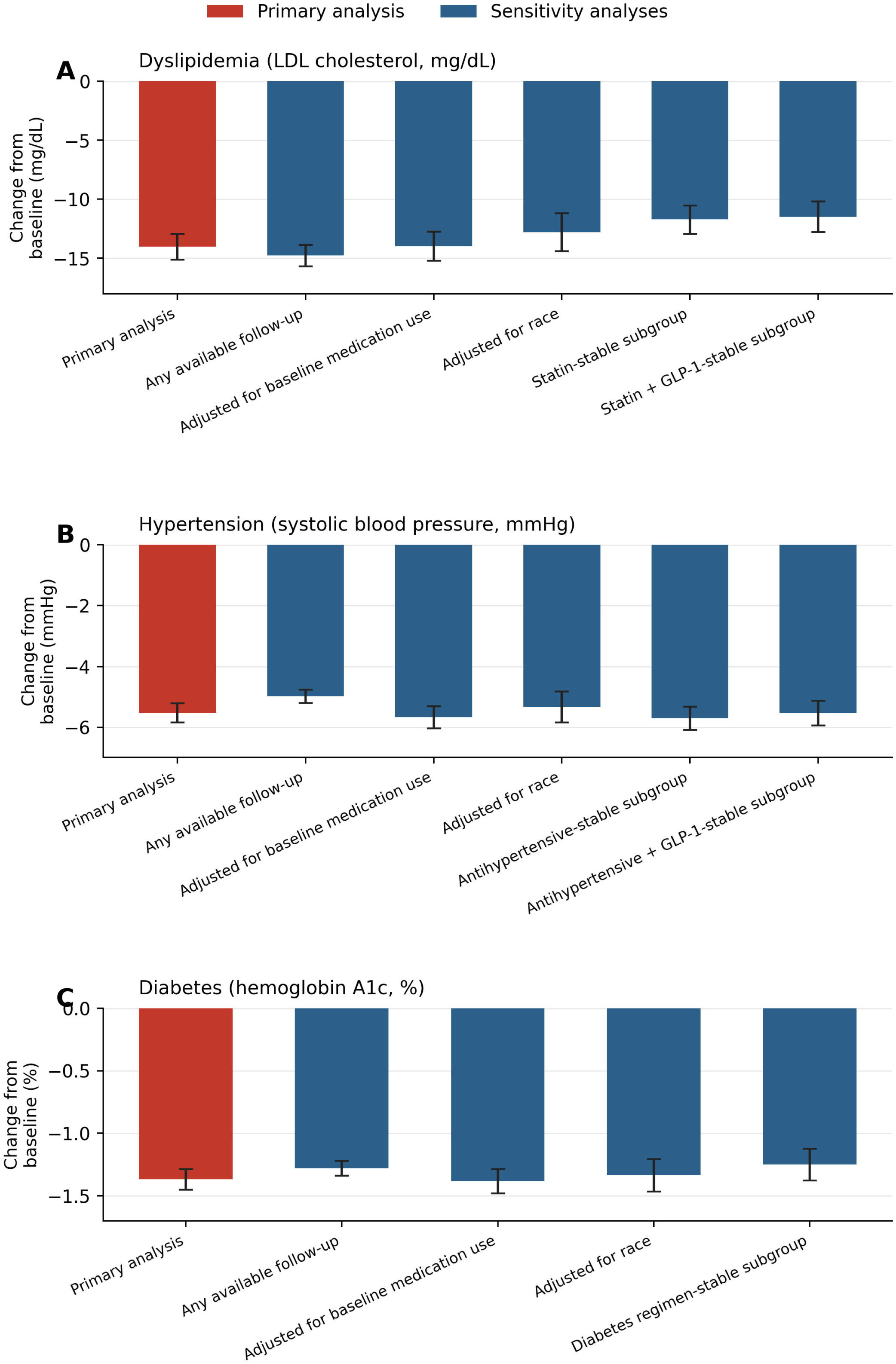
Cardiometabolic Improvements at 6 Months Were Consistent Across Primary and Sensitivity Analyses. Six-month change from baseline in low-density lipoprotein (LDL) cholesterol (Panel A; primary analysis n=6,347), systolic blood pressure (Panel B; primary analysis n=13,573), and hemoglobin A1c (Panel C; primary analysis n=2,394), estimated as the model-based estimated marginal mean (EMM) from linear mixed models comparing the 180-day to the baseline value. Red bars indicate the primary analysis; blue bars indicate sensitivity analyses testing the robustness of the primary specification to follow-up completeness, unmeasured covariate adjustment, and medication changes. “Any available follow-up” re-estimates the primary model among all patients with any available follow-up value, regardless of whether there is independent evidence they remained actively engaged in care. “Adjusted for baseline medication use” and “adjusted for race” add that covariate to the primary model, restricted to patients with complete data for it (sample sizes for all specifications are reported in the tables). Subgroup analyses restrict to patients whose relevant medication regimen (statin, antihypertensive, or diabetes regimen, with or without a glucagon-like peptide-1 [GLP-1] receptor agonist) was unchanged from baseline through 6 months. Error bars represent 95% confidence intervals, calculated from the exact model-based pairwise contrast (6 months vs baseline) for each specification.

### Intervention

Nourish delivers vMNT through a telehealth platform in which an RD serves as the primary clinician. Care was delivered by licensed RDs guided by condition-specific, evidence-based clinical care pathways that provide recommended frameworks for phased care delivery and dietary intervention. RDs drew on pathway recommendations, clinical judgment, and patient preferences to develop individualized treatment plans. Pathway recommendations emphasized dietary fat quality and fiber intake for dyslipidemia, DASH-aligned dietary approaches for hypertension, and meal structure, carbohydrate quality, and composition to attenuate postprandial glucose excursions for diabetes. Sessions focused on individualized dietary assessment, goal setting, and ongoing nutrition counseling, with visit cadence determined by clinical need and engagement. Between visits, patients could log dietary intake and other health data, message their RD directly, access educational materials, and receive AI-generated suggestions and feedback through the Nourish app. Care was delivered as usual care. Coverage was primarily through major insurance plans, with a subset of patients paying out of pocket.

### Data Sources

Clinical outcome data, including laboratory values, blood pressure, and weight, were collected as part of usual care and obtained from the Nourish EHR, which aggregates data from multiple sources including health information exchange connections, patient-reported values entered during visits or via the Nourish app, and connected devices and mobile apps. Medication data were obtained from pharmacy records via health information exchange; patients with any available pharmacy data were assumed to have complete medication records, and this subset was used exclusively for all medication analyses. Demographic data were obtained from patient intake, including age, sex, race, and ethnicity self-reported at enrollment, and BMI computed from self-reported height and weight recorded at the first RD visit.

### Statistical Analysis

Outcomes were analyzed using linear mixed models with maximum likelihood estimation and an unstructured covariance structure, with time modeled as a fixed effect at enrollment, 3, and 6 months. All models were adjusted for age, sex, and BMI at enrollment. Medication use and race were excluded as covariates from primary models due to missing data in approximately 22–25% and 51–59% of patients, respectively; sensitivity analyses were performed adding these variables as covariates restricted to patients with available data. Pairwise comparisons were used to estimate within-period change and associated standard errors at each timepoint. Outcome values were assigned to the nearest timepoint within predefined measurement windows.

Maximum likelihood estimation accommodates missing outcome data under the missing-at-random assumption, retaining all patients with at least one outcome observation in the model. To assess potential selection bias from the outcome requirement, we compared baseline characteristics between patients with and without 6-month outcome data within the engaged cohort; those with follow-up data were more likely to have prevalent diabetes and GLP-1 use, consistent with more intensive monitoring in these groups, while other characteristics were similar between groups, broadly supporting the missing-at-random assumption. A sensitivity analysis using a cohort defined by outcome data availability alone, without the engagement requirement, was also performed.

To characterize medication use during the study period and examine outcomes independent of medication change, we conducted pre-specified analyses among patients with available pharmacy data. We first used McNemar’s tests to assess within-patient change in medication use from enrollment to 6 months. Drug classes examined were cohort-specific: statins, fibrates, and glucagon-like-peptide-1 (GLP-1) receptor agonists (dyslipidemia); antihypertensives and GLP-1 agonists (hypertension); and GLP-1 agonists, metformin, insulin, sodium-glucose cotransporter-2 (SGLT-2) inhibitors, sulfonylureas, and dipeptidyl peptidase-4 (DPP-4) inhibitors (diabetes). For medication classes that increased significantly during the study period, we conducted subgroup analyses among patients with stable use of those classes (no new initiations or discontinuations across all three timepoints) to characterize outcomes independent of medication intensification.

We additionally evaluated the proportion of patients presenting above guideline-recommended targets who had 6-month outcome data available and achieved those targets by 6 months: HbA1c less than 7.0% per American Diabetes Association (ADA) recommendations, and SBP less than 130 mmHg per American Heart Association (AHA) and American College of Cardiology (ACC) guidelines.

All analyses were conducted using IBM SPSS Statistics version 32; statistical significance was set at p<0.05.

## Results

### Population Characteristics

Enrollment characteristics are summarized in Table 1. Across cohorts, patients were predominantly female and with a mean (SD) age of 42.9 (11.6) and BMI of 35.5 (8.6). Patients resided in all 50 states and the District of Columbia, with the largest proportions from the South. Among patients with available race data, the majority identified as White, although race data were missing in more than half of patients in each cohort. Adults with diabetes were somewhat older (mean, 48.0 [12.6] years) with higher BMI at enrollment compared with the other cohorts. At baseline, mean (SD) HbA1c was 8.1 (1.8) % in the diabetes cohort, SBP was 130.5 ( 12.9) mmHg in the hypertension cohort, and LDL-C was 135.4 (36.1) mg/dL in the dyslipidemia cohort.

**Table 1.**
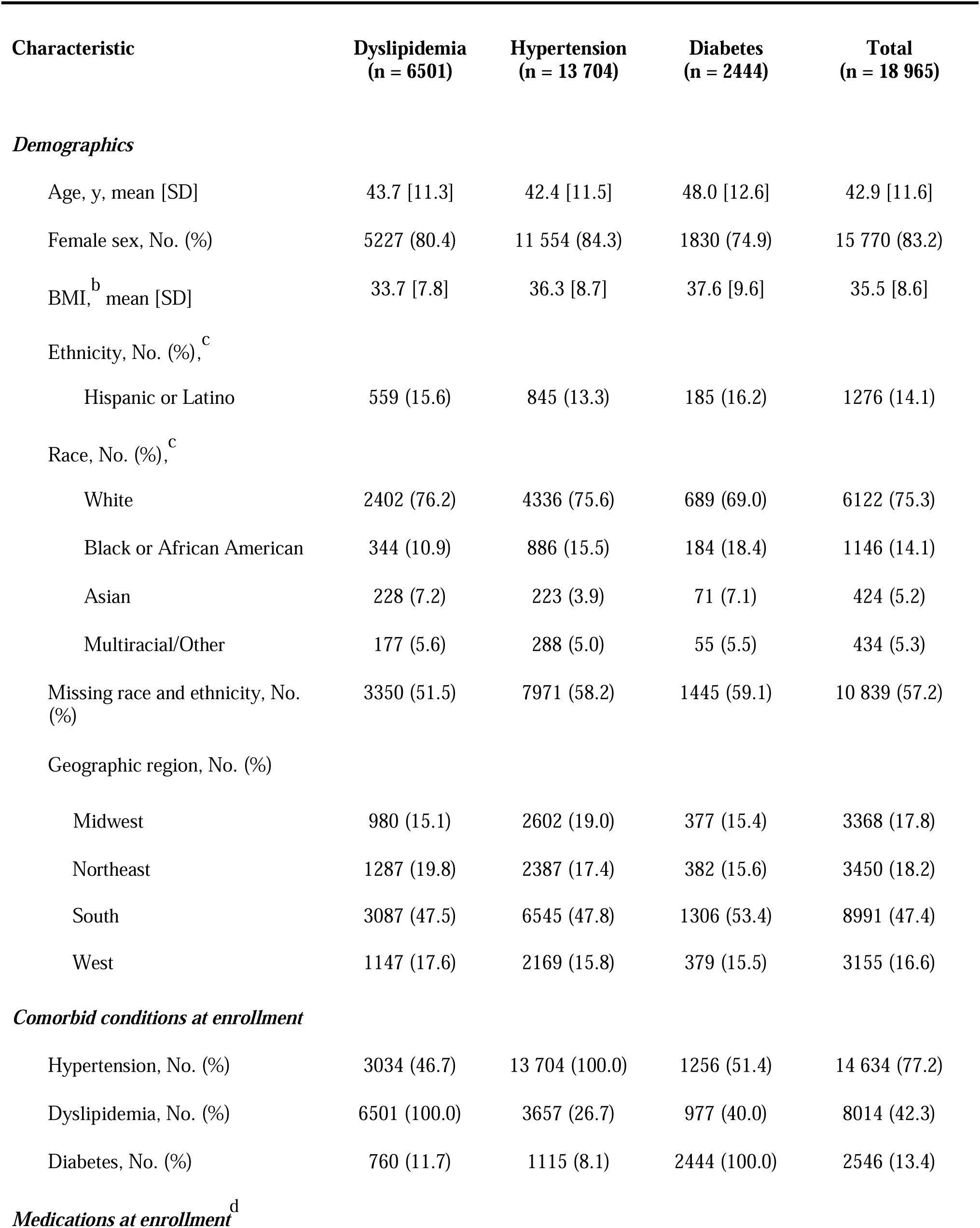
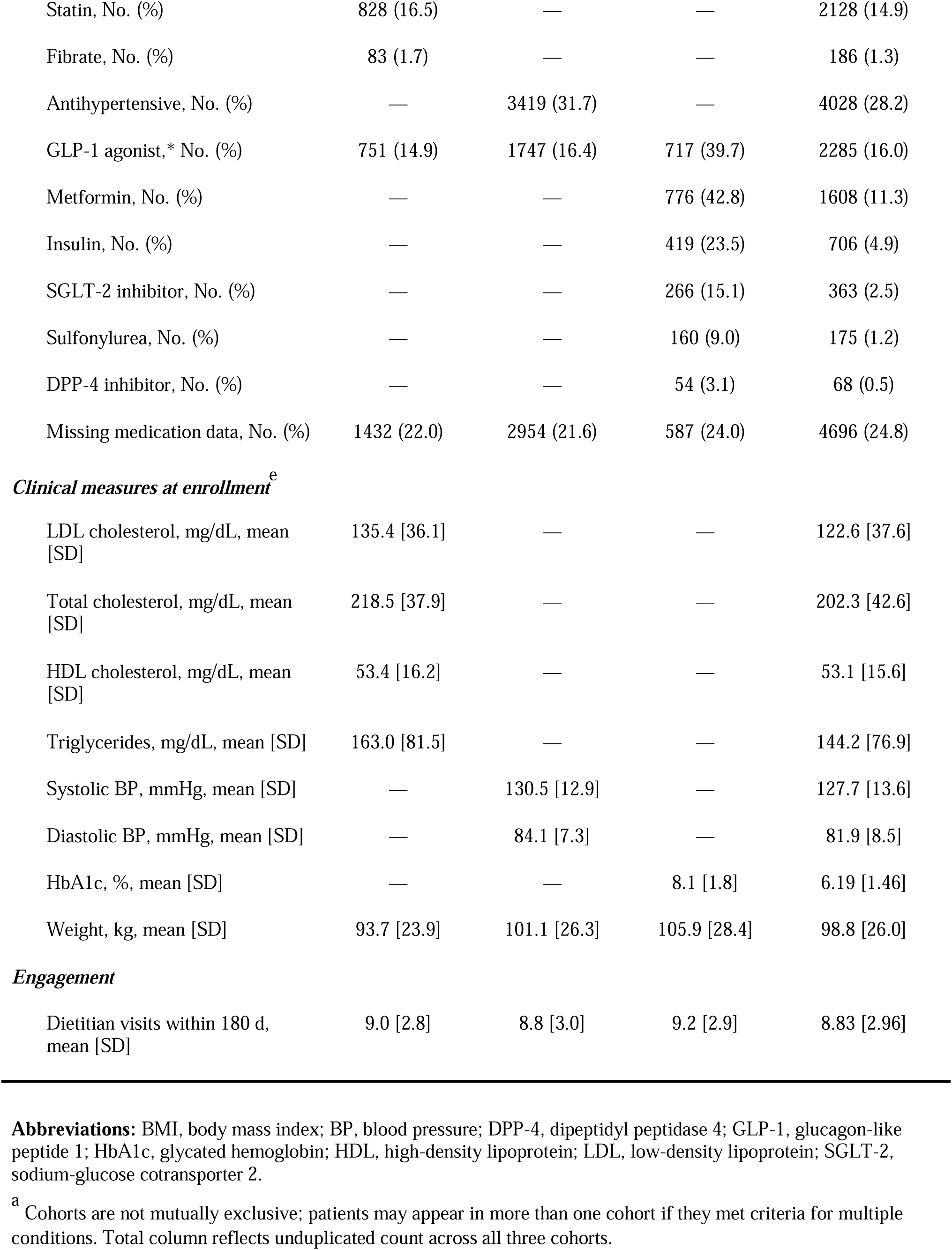
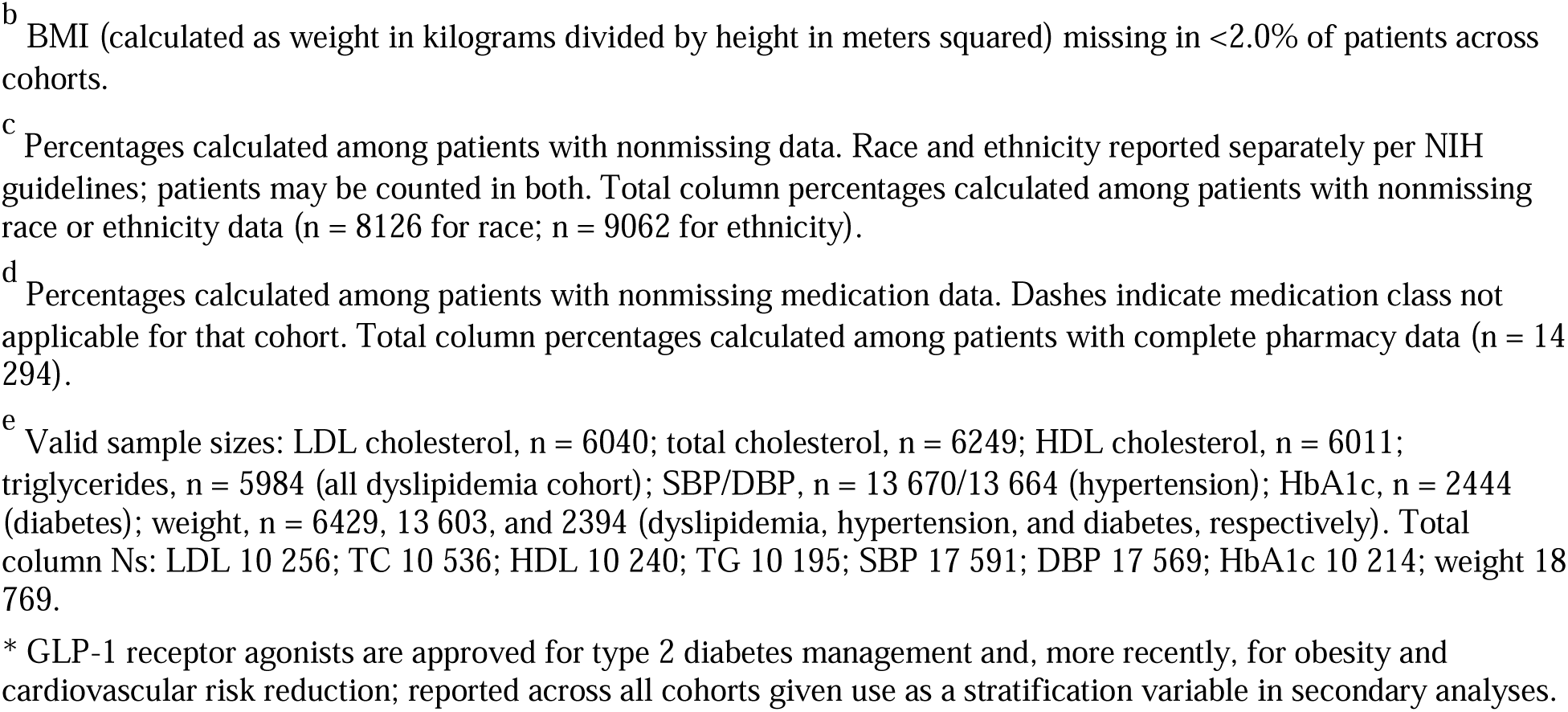
Enrollment characteristics by cohort.

### Primary Outcomes

#### Dyslipidemia

Among individuals with dyslipidemia at baseline, levels of all four lipid markers declined over time (p<0.0001 each; Table 2). Compared with baseline, LDL-C and triglycerides were 12.8 and 26.4 mg/dl lower at 3 months, and 14.0 and 28.6 mg/dl lower at 6 months, respectively, with LDL-C continuing to decline modestly between 3 and 6 months (p=.036). HDL-C showed a small but statistically significant decline of -0.5 mg/dl at 6 months. Weight in this cohort declined by 3.8 kg (−4.0%) at 6 months (p<0.001).

**Table 2.** Primary linear mixed model results: estimated marginal means and changes from baseline^a,c^.

| Outcome | Baseline value<br>[SE] | 3 mo |  | 6mo |  |
| --- | --- | --- | --- | --- | --- |
|  |  | Value [SE] | Change [SE]<sup>b</sup> | Value [SE] | Change [SE]<sup>b</sup> |
| Dyslipidemia cohort (n = 6501) |  |  |  |  |  |
| LDL cholesterol, mg/dL | 135.2 [0.46] | 122.5 [0.53] | −12.8 [0.53]*** | 121.2 [0.54] | −14.0 [0.56]*** |
| Total cholesterol, mg/dL | 218.2 [0.47] | 199.9 [0.59] | −18.4 [0.60]*** | 198.9 [0.60] | −19.4 [0.61]*** |
| HDL cholesterol, mg/dL | 53.4 [0.19] | 52.2 [0.19] | −1.2 [0.15]*** | 52.9 [0.19] | −0.5 [0.15]** |
| Triglycerides, mg/dL | 162.8 [1.02] | 136.4 [0.99] | −26.4 [1.00]*** | 134.2 [1.04] | −28.6 [1.06]*** |
| Weight, kg | 94.3 [0.11] | 91.9 [0.12] | −2.4 [0.07]*** | 90.6 [0.14] | −3.8 [0.10]*** |
| Hypertension cohort (n = 13 704) |  |  |  |  |  |
| Systolic BP, mmHg | 130.5 [0.11] | 125.3 [0.13] | −5.3 [0.15]*** | 125.0 [0.14] | −5.5 [0.16]*** |
| Diastolic BP, mmHg | 84.1 [0.06] | 79.7 [0.09] | −4.4 [0.10]*** | 79.6 [0.10] | −4.5 [0.11]*** |
| Weight, kg | 101.4 [0.08] | 99.3 [0.09] | −2.1 [0.05]*** | 98.0 [0.10] | −3.5 [0.07]*** |
| Diabetes cohort (n = 2444) |  |  |  |  |  |
| HbA1c, % | 8.07 [0.04] | 6.92 [0.03] | −1.15 [0.04]*** | 6.70 [0.03] | −1.37 [0.04]*** |
| Weight, kg | 106.4 [0.20] | 103.1 [0.24] | −3.3 [0.14]*** | 101.2 [0.26] | −5.2 [0.19]*** |

**Abbreviations:** BP, blood pressure; HbA1c, glycated hemoglobin; HDL, high-density lipoprotein; LDL, low-density lipoprotein; SE, standard error.
<sup>a</sup> All models adjusted for age, sex, and BMI at enrollment. Values are estimated marginal means. Estimated using linear mixed models with maximum likelihood estimation and unstructured covariance. Weight is a secondary outcome reported across all cohorts.
<sup>b</sup> Change from baseline. \*\*\* $p < 0.001$ ; \*\* $p < 0.01$ ; \* $p < 0.05$ . HbA1c and weight continued to decline significantly from 3 to 6 months across all cohorts (all $p < .001$ ). Systolic BP, diastolic BP, and most lipid outcomes plateaued from 90 to 180 days ( $p > .05$ ), with the exception of LDL cholesterol, which continued to decline modestly ( $p = 0.036$ ).
<sup>c</sup> Sample sizes varied by outcome due to differing data availability and covariate completeness. Dyslipidemia cohort: LDL cholesterol, $n = 6347$ ; total cholesterol, $n = 6397$ ; HDL cholesterol, $n = 6357$ ; triglycerides, $n = 6354$ ; weight, $n = 6418$ . Hypertension cohort: systolic BP, $n = 13\,573$ ; diastolic BP, $n = 13\,573$ ; weight, $n = 13\,574$ . Diabetes cohort: HbA1c, $n = 2394$ ; weight, $n = 2394$ .

#### Hypertension

Among those with elevated BP, SBP and DBP declined by 5.3 and 4.4 mmHg, respectively, by 3 months, with these improvements maintained through 6 months (SBP −5.5 mmHg, DBP −4.5 mmHg; p<0.0001 each; Table 2). Among the 4,504 patients presenting with SBP ≥130 mmHg at enrollment who also had 6-month SBP data available, 2579 (57.3%) achieved the AHA/ACC target of less than 130 mmHg by 6 months. Weight declined by 3.5 kg (−3.4%) at 6 months.

#### Diabetes

Among individuals with diabetes, HbA1c declined significantly by 3 months and continued to decline through 6 months, with a mean total reduction of 1.37% from enrollment to 6 months (p<0.0001 across timepoints; Table 2). Among the 1,106 patients presenting with HbA1c ≥7.0% at enrollment who had 6-month HbA1c data available, 605 (54.7%) achieved the ADA target of less than 7.0% by 6 months. Weight declined by 5.2 kg (−4.9%) at 6 months.

### Medication Use During the Study Period

Medication use changed significantly during the study period across all three cohorts (Table 3). GLP-1 agonist use increased across all cohorts, rising by 8.2, 7.9, and 10.5 percentage points in the dyslipidemia, hypertension, and diabetes cohorts, respectively (all p<0.001). Statin use increased modestly in the dyslipidemia cohort (+2.8 percentage points, p<0.001); fibrate use did not change. Antihypertensive use increased modestly in the hypertension cohort (+3.0 percentage points, p<.001). In the diabetes cohort, insulin and SGLT-2 inhibitor use increased modestly (+3.1 and +2.5 percentage points, respectively; both p<.001), while metformin, sulfonylurea, and DPP-4 inhibitor use did not change.

**Table 3.** Medication use at enrollment and 6 months by cohort^a^.

| Medication | Enrollment,<br>No. (%) | 6 mo,<br>No. (%) | Net change,<br>No. (%) | P value |
| --- | --- | --- | --- | --- |
| **Dyslipidemia cohort (medication data n = 4980–5033)<sup>b</sup>** |  |  |  |  |
| Statin, No. (%) | 828 (16.5) | 971 (19.3) | 143 (2.8) | <0.001 |
| Fibrate, No. (%) | 83 (1.7) | 94 (1.9) | 11 (0.2) | 0.177 |
| GLP-1 agonist, No. (%)<sup>*</sup> | 747 (14.9) | 1159 (23.1) | 412 (8.2) | <0.001 |
| **Hypertension cohort (medication data n = 10 625–10 767)<sup>c</sup>** |  |  |  |  |
| Antihypertensive, No. (%) | 3419 (31.7) | 3733 (34.7) | 322 (3.0) | <0.001 |
| GLP-1 agonist, No. (%)<sup>*</sup> | 1744 (16.4) | 2587 (24.3) | 843 (7.9) | <0.001 |
| **Diabetes cohort (medication data n = 1756–1810)<sup>d</sup>** |  |  |  |  |
| GLP-1 agonist, No. (%)<sup>*</sup> | 714 (39.6) | 904 (50.2) | 190 (10.5) | <0.001 |
| Metformin, No. (%) | 773 (42.7) | 773 (42.7) | 0 (0.0) | 1.000 |
| Insulin, No. (%) | 419 (23.5) | 475 (26.6) | 56 (3.1) | <0.001 |
| SGLT-2 inhibitor, No. (%) | 266 (15.1) | 311 (17.6) | 45 (2.5) | <0.001 |
| Sulfonylurea, No. (%) | 159 (9.0) | 148 (8.4) | –11 (–0.6) | 0.305 |
| DPP-4 inhibitor, No. (%) | 54 (3.1) | 44 (2.5) | –10 (–0.6) | 0.143 |

**Abbreviations:** DPP-4, dipeptidyl peptidase 4; GLP-1, glucagon-like peptide 1; SGLT-2, sodium-glucose cotransporter 2.
<sup>a</sup> Percentages calculated among patients with nonmissing medication data at both enrollment and 6 months. McNemar test for paired proportions assesses within-patient change. Variation in valid N across medications within cohort reflects a small number of patients (<2%) with partial medication data; missingness is otherwise person-level.
<sup>b</sup> Full dyslipidemia cohort n = 6501; medication data missing in 22.0%–23.4% of patients.
<sup>c</sup> Full hypertension cohort n = 13 704; medication data missing in 21.6%–22.5% of patients.
<sup>d</sup> Full diabetes cohort n = 2444; medication data missing in 26.0%–28.2% of patients.

Given these changes, we conducted pre-specified subgroup analyses among patients with stable medication regimens to characterize outcomes independent of medication change. Across all cohorts, patients with stable regimens showed meaningful improvements in primary outcomes and weight, consistent in magnitude with the full cohort (Table 4); for example, the diabetes cohort’s fully stable subgroup (n=891) showed a mean HbA1c reduction of 1.25% and weight loss of 4.9 kg.

**Table 4.**
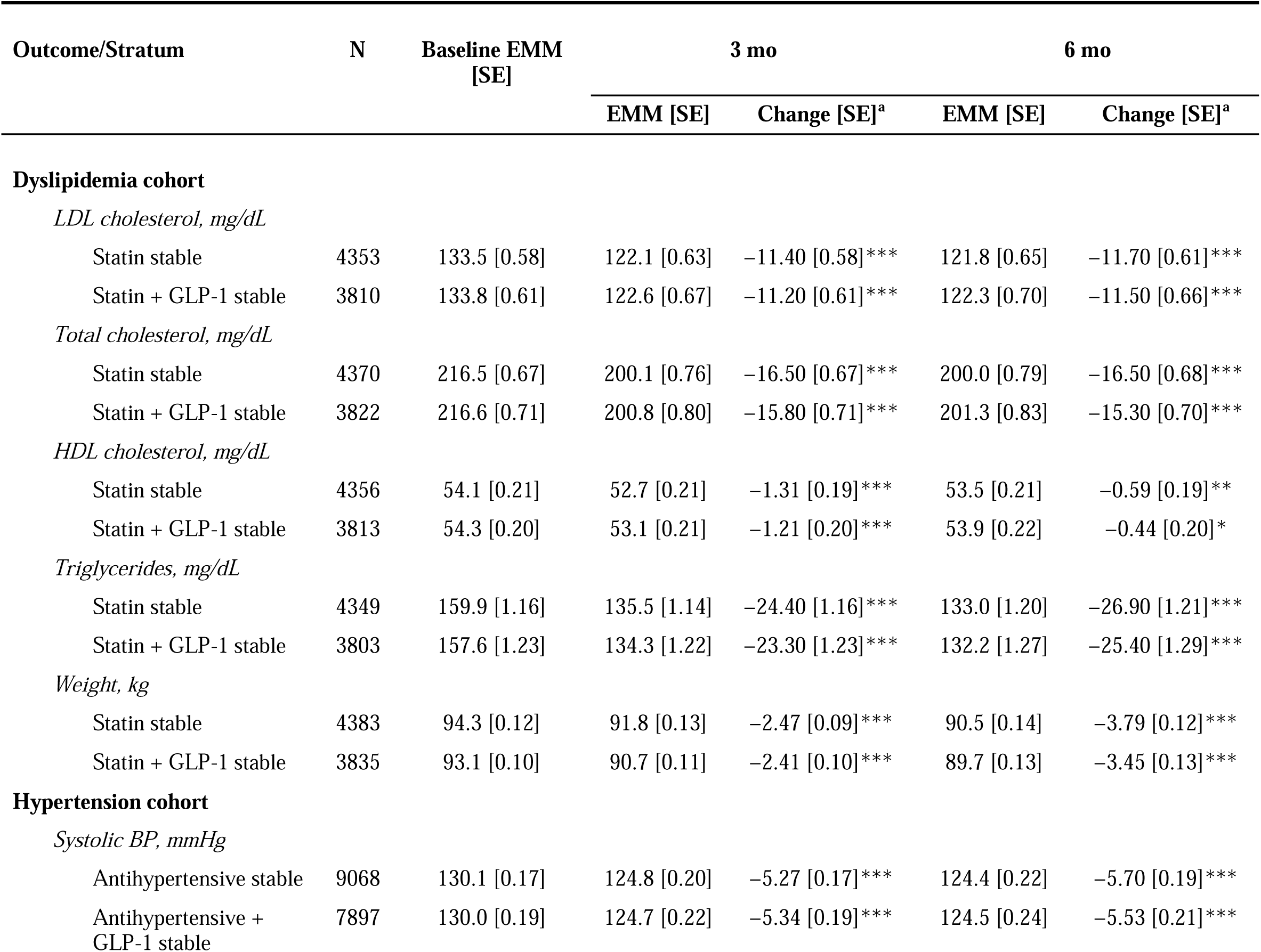
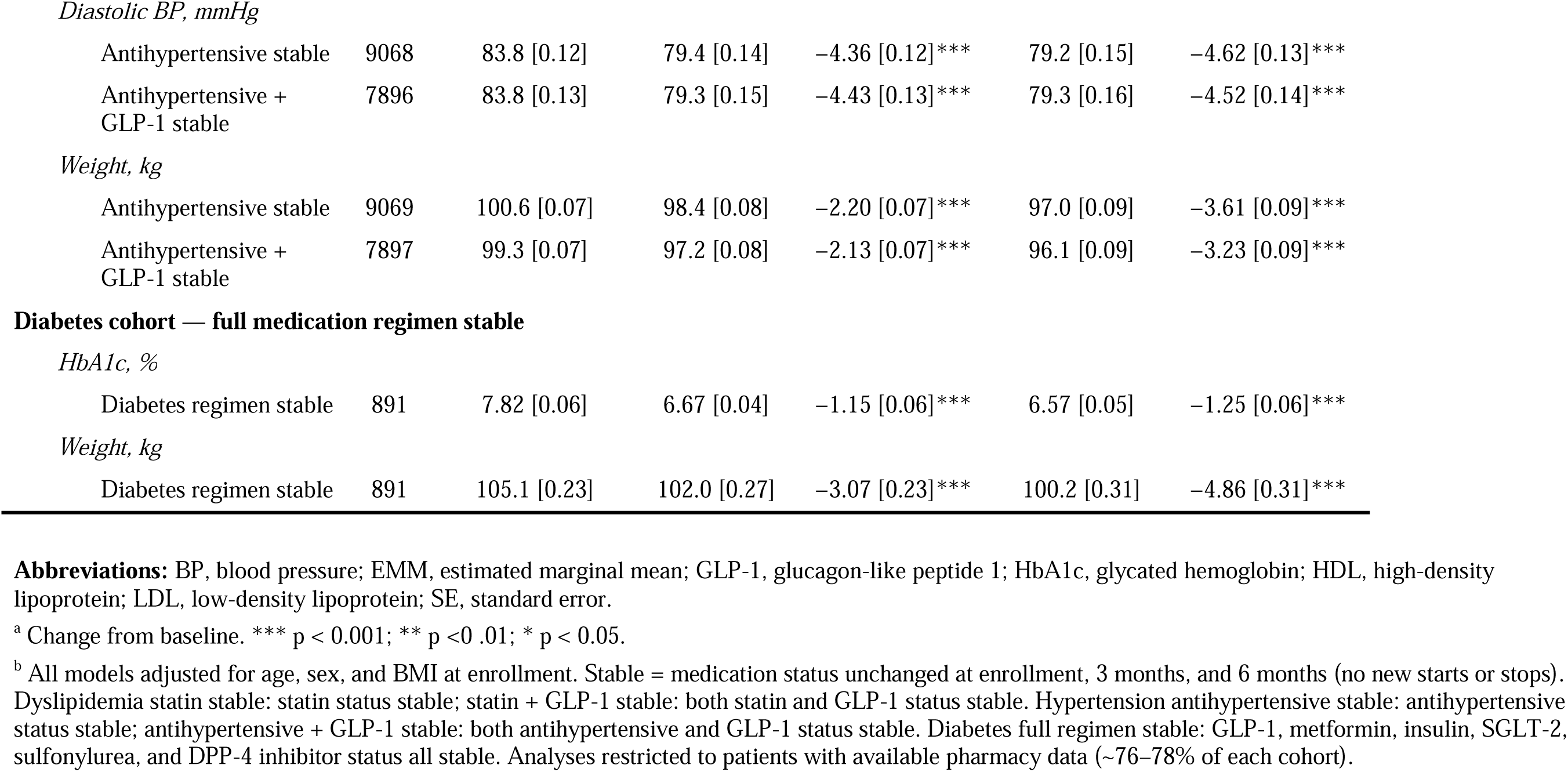
Subgroup analyses by medication stability^b^.

| Outcome/Stratum | N | Baseline EMM<br>[SE] | 3 mo |  | 6 mo |  |
| --- | --- | --- | --- | --- | --- | --- |
|  |  |  | EMM [SE] | Change [SE]<sup>a</sup> | EMM [SE] | Change [SE]<sup>a</sup> |
| Dyslipidemia cohort |  |  |  |  |  |  |
| LDL cholesterol, mg/dL |  |  |  |  |  |  |
| Statin stable | 4353 | 133.5 [0.58] | 122.1 [0.63] | −11.40 [0.58]*** | 121.8 [0.65] | −11.70 [0.61]*** |
| Statin + GLP-1 stable | 3810 | 133.8 [0.61] | 122.6 [0.67] | −11.20 [0.61]*** | 122.3 [0.70] | −11.50 [0.66]*** |
| Total cholesterol, mg/dL |  |  |  |  |  |  |
| Statin stable | 4370 | 216.5 [0.67] | 200.1 [0.76] | −16.50 [0.67]*** | 200.0 [0.79] | −16.50 [0.68]*** |
| Statin + GLP-1 stable | 3822 | 216.6 [0.71] | 200.8 [0.80] | −15.80 [0.71]*** | 201.3 [0.83] | −15.30 [0.70]*** |
| HDL cholesterol, mg/dL |  |  |  |  |  |  |
| Statin stable | 4356 | 54.1 [0.21] | 52.7 [0.21] | −1.31 [0.19]*** | 53.5 [0.21] | −0.59 [0.19]** |
| Statin + GLP-1 stable | 3813 | 54.3 [0.20] | 53.1 [0.21] | −1.21 [0.20]*** | 53.9 [0.22] | −0.44 [0.20]* |
| Triglycerides, mg/dL |  |  |  |  |  |  |
| Statin stable | 4349 | 159.9 [1.16] | 135.5 [1.14] | −24.40 [1.16]*** | 133.0 [1.20] | −26.90 [1.21]*** |
| Statin + GLP-1 stable | 3803 | 157.6 [1.23] | 134.3 [1.22] | −23.30 [1.23]*** | 132.2 [1.27] | −25.40 [1.29]*** |
| Weight, kg |  |  |  |  |  |  |
| Statin stable | 4383 | 94.3 [0.12] | 91.8 [0.13] | −2.47 [0.09]*** | 90.5 [0.14] | −3.79 [0.12]*** |
| Statin + GLP-1 stable | 3835 | 93.1 [0.10] | 90.7 [0.11] | −2.41 [0.10]*** | 89.7 [0.13] | −3.45 [0.13]*** |
| Hypertension cohort |  |  |  |  |  |  |
| Systolic BP, mmHg |  |  |  |  |  |  |
| Antihypertensive stable | 9068 | 130.1 [0.17] | 124.8 [0.20] | −5.27 [0.17]*** | 124.4 [0.22] | −5.70 [0.19]*** |
| Antihypertensive +<br>GLP-1 stable | 7897 | 130.0 [0.19] | 124.7 [0.22] | −5.34 [0.19]*** | 124.5 [0.24] | −5.53 [0.21]*** |
| *Diastolic BP, mmHg* |  |  |  |  |  |  |
| Antihypertensive stable | 9068 | 83.8 [0.12] | 79.4 [0.14] | −4.36 [0.12]*** | 79.2 [0.15] | −4.62 [0.13]*** |
| Antihypertensive +<br>GLP-1 stable | 7896 | 83.8 [0.13] | 79.3 [0.15] | −4.43 [0.13]*** | 79.3 [0.16] | −4.52 [0.14]*** |
| *Weight, kg* |  |  |  |  |  |  |
| Antihypertensive stable | 9069 | 100.6 [0.07] | 98.4 [0.08] | −2.20 [0.07]*** | 97.0 [0.09] | −3.61 [0.09]*** |
| Antihypertensive +<br>GLP-1 stable | 7897 | 99.3 [0.07] | 97.2 [0.08] | −2.13 [0.07]*** | 96.1 [0.09] | −3.23 [0.09]*** |
| **Diabetes cohort — full medication regimen stable***HbA1c, %* |  |  |  |  |  |  |
| Diabetes regimen stable | 891 | 7.82 [0.06] | 6.67 [0.04] | −1.15 [0.06]*** | 6.57 [0.05] | −1.25 [0.06]*** |
| *Weight, kg* |  |  |  |  |  |  |
| Diabetes regimen stable | 891 | 105.1 [0.23] | 102.0 [0.27] | −3.07 [0.23]*** | 100.2 [0.31] | −4.86 [0.31]*** |

**Abbreviations:** BP, blood pressure; EMM, estimated marginal mean; GLP-1, glucagon-like peptide 1; HbA1c, glycated hemoglobin; HDL, high-density lipoprotein; LDL, low-density lipoprotein; SE, standard error.
<sup>a</sup> Change from baseline. \*\*\* p < 0.001; \*\* p < 0.01; \* p < 0.05.
<sup>b</sup> All models adjusted for age, sex, and BMI at enrollment. Stable = medication status unchanged at enrollment, 3 months, and 6 months (no new starts or stops). Dyslipidemia statin stable: statin status stable; statin + GLP-1 stable: both statin and GLP-1 status stable. Hypertension antihypertensive stable: antihypertensive status stable; antihypertensive + GLP-1 stable: both antihypertensive and GLP-1 status stable. Diabetes full regimen stable: GLP-1, metformin, insulin, SGLT-2, sulfonyleurea, and DPP-4 inhibitor status all stable. Analyses restricted to patients with available pharmacy data (~76–78% of each cohort).

### Sensitivity Analyses

Sensitivity analyses evaluated all patients with outcome data regardless of engagement duration, adjusted for baseline medication use, and adjusted for race. In each, results were consistent with primary findings across all cohorts (eTables 1–3; Figure 1).

## Discussion

In this large, real-world cohort of nearly 19,000 patients across all 50 states and the District of Columbia, vMNT delivered through a technology-enabled care model was associated with consistent, clinically meaningful improvements across dyslipidemia, hypertension, and diabetes — three core metabolic risk factors of cardiovascular-kidney-metabolic syndrome.^1^ To our knowledge, this is among the first studies to characterize real-world vMNT outcomes at this scale across multiple related cardiometabolic conditions, extending prior evidence on MNT to a technology-supported virtual care setting.

Among adults with dyslipidemia, mean LDL-C, total cholesterol, and triglycerides each declined to below standard clinical thresholds by 6 months. LDL-C and triglycerides showed the largest, most sustained improvements, with LDL continuing to decline modestly between 3 and 6 months. HDL-C declined by 0.5 mg/dL, a small but statistically significant change of unclear cause that was outweighed by the larger, favorable changes in LDL-C and triglycerides. These reductions are clinically meaningful: LDL-C lowering reduces major cardiovascular events in large randomized trials, with risk reduction roughly proportional to the absolute reduction achieved,^14^ and triglyceride-rich remnant lipoproteins have similarly been implicated as a causal contributor to residual cardiovascular risk beyond LDL-C, with triglyceride-lowering associated with reduced cardiovascular events across multiple therapeutic approaches.^15^ The magnitudes of these reductions are consistent with the causal, between-group effects reported in a meta-analysis of 7 small randomized controlled trials (RCTs) of dietitian-delivered MNT (n=838), primarily delivered in person among patients with hypercholesterolemia or other specific lipid abnormalities: total cholesterol was reduced by 20.8 mg/dL (compared with 19.4 in our study), LDL-C by 11.6 mg/dl (compared with 14.0 in our study), and triglycerides by 32.6 mg/dl (compared with 28.6 mg/dl in our study).^16^ While our results are observational, this consistency with RCT effects provides some reassurance and biological plausibility that the present findings are not entirely due to bias or regression toward the mean. This study extends that prior evidence to a broader, real-world population with MNT delivered virtually at scale.

More than half of the individuals not meeting BP targets at enrollment achieved the AHA/ACC target of less than 130 mmHg by 6 months (57.3%), with mean SBP declining by 5.5 mmHg and remaining stable from 3 to 6 months. A reduction of this magnitude is clinically meaningful: each 2 mmHg difference in usual SBP has been associated with ∼7% and 10% differences in mortality from ischemic heart disease and stroke, respectively, in a meta-analysis of over 1 million adults,^17^ and RCTs have further demonstrated that intervention-driven reductions reduce cardiovascular events and mortality.^18^ In a meta-analysis of 31 randomized trials of dietitian-delivered MNT, nearly all in-person or blended, MNT reduced SBP by 3.6 mmHg and DBP by 2.0 mmHg among patients with hypertension or prehypertension.^19^ As with dyslipidemia, our results in a far larger, more diverse sample treated with vMNT extend these findings with contemporary evidence on changes in BP levels.

In our cohort, more than half of patients presenting with HbA1c above the ADA-recommended target of 7.0% achieved that target by 6 months (54.7%), with mean HbA1c declining by 1.37 percentage points. Each 1% reduction in HbA1c has been associated with a 21% lower risk of diabetes-related complications in large observational studies.^20^ Our 1.37-point reduction substantially exceeded both the causal, between-group effect (0.47 percentage points) and the pre-post change within intervention arms (0.65 percentage points) reported in a meta-analysis of 13 RCTs (2,511 subjects) of dietitian-delivered dietetic intervention in diabetes, with durations ranging from 4 months to 3 years.^21^ Whether these findings relate to more effective modern glucose-focused nutrition recommendations emphasizing healthy fats, fiber, and protein over refined carbohydrates, or greater efficacy of vMNT’s technology-enabled support between visits, cannot be differentiated.

Weight declined meaningfully across all three cohorts and continued to decline through 6 months. Weight loss is known to improve blood lipids, BP, and glycemic control. However, the magnitude of improvement observed here exceeds what would typically be expected from weight loss of this amount alone, based on dose-response relationships reported in general populations^22,23^ and in diabetes,^24,25^ suggesting improved diet quality and composition also contributed independently. Controlled feeding trials support this interpretation: weight-stable dietary interventions varying food composition can substantially improve BP, lipid profiles, and glycemic control and insulin sensitivity, independent of weight change.^26–28^

Medications intended to treat these conditions, including GLP-1 agonists, were used at baseline and, in some cases, added during the study period across all three cohorts. Among patients with stable medication regimens (no initiations or discontinuations across all three timepoints), outcomes were of similar or only modestly smaller magnitude than in the full cohort. This suggests MNT may further contribute to improvements beyond concurrent pharmacotherapy, particularly relevant given rising GLP-1 utilization nationally.^29^

This study has several strengths. The large sample size increased the precision of these estimates and enabled well-powered subgroup analyses, including the medication-stability analyses, within a population receiving usual clinical care. The cohort also spanned all 50 states and the District of Columbia, demonstrating this model’s capacity to deliver MNT at scale to populations who may otherwise lack local access to in-person care. Finally, the technology-enabled model may have contributed to the magnitude of improvement observed, though this cannot be established directly from the present design.

This study also has limitations. The absence of a control group precludes ruling out regression to the mean or secular trends, so causality cannot be established; nonetheless, the consistency of these reductions with those reported in RCTs of dietitian-delivered MNT makes regression or bias unlikely to fully explain the results. As with other real-world data sources, this study was subject to missing data and potential selection bias, including incomplete capture of medication use and race, and reliance on a cohort with available follow-up outcome data; sensitivity analyses addressing these gaps were consistent with primary findings, supporting confidence in the robustness of these results. This single-arm design cannot isolate the independent contribution of the combined care model’s components, including the RD counseling and technology-enabled support. Future studies should incorporate comparison groups, extend follow-up beyond 6 months to assess durability, and identify which model components drive successful outcomes.

In this large, real-world study, vMNT delivered through a technology-enabled care model was associated with clinically meaningful improvement across three prevalent cardiometabolic conditions. These findings position technology-enabled virtual care as a promising approach for delivering effective MNT at scale, particularly for populations with limited access to in-person care.

## Supporting information

eSupplement

eFigure 1

## Data Availability

Deidentified individual patient data are not available, as they are proprietary to Nourish, Inc, and subject to data use restrictions.

## Author Contributions

Dr. Adams had full access to all the data in the study and takes responsibility for the integrity of the data and the accuracy of the data analysis.

*Concept and design*: Adams, Mozaffarian. *Acquisition, analysis, or interpretation of data:* Adams, Bollinger, Mozaffarian. *Drafting of the manuscript:* Adams. *Critical revision of the manuscript for important intellectual content:* Adams, Bollinger, Mozaffarian. *Statistical analysis:* Adams. *Supervision:* Mozaffarian.

## Additional Contributions

The authors thank the Nourish registered dietitians and clinical care team who provided care to the patients described in this study.

## Use of Artificial Intelligence

Claude (Anthropic) was used to assist with drafting, editing, and formatting portions of this manuscript, including figure and table construction, between June 2026 and September 2026; multiple model versions were used over this period. The authors take full responsibility for the integrity of the content generated with this assistance.

