## Supplementary material for "Real-World Outcomes of Virtual Medical Nutrition Therapy in Dyslipidemia, Hypertension, or Diabetes": eSupplement

**eTable 1.** Sensitivity analysis: primary models with baseline medication covariates^b^

| **Outcome** | **N** | **Baseline EMM [SE]** | **90 d** | | **180 d** | |
| --- | --- | --- | --- | --- | --- | --- |
|  |  |  | **EMM [SE]** | **Change [SE]^a^** | **EMM [SE]** | **Change [SE]^a^** |
| **Dyslipidemia cohort** | | | | | | |
| LDL cholesterol, mg/dL | 4908 | 134.3 [0.52] | 121.8 [0.56] | −12.52 [0.59]*** | 120.3 [0.59] | −13.98 [0.63]*** |
| Total cholesterol, mg/dL | 4930 | 217.4 [0.54] | 199.4 [0.64] | −18.01 [0.67]*** | 198.1 [0.67] | −19.27 [0.69]*** |
| HDL cholesterol, mg/dL | 4909 | 53.7 [0.21] | 52.5 [0.22] | −1.22 [0.18]*** | 53.2 [0.21] | −0.55 [0.18]** |
| Triglycerides, mg/dL | 4898 | 161.3 [1.13] | 135.8 [1.11] | −25.45 [1.10]*** | 133.5 [1.17] | −27.77 [1.18]*** |
| Weight, kg | 4943 | 94.3 [0.12] | 91.8 [0.14] | −2.46 [0.09]*** | 90.5 [0.16] | −3.81 [0.11]*** |
| **Hypertension cohort** | | | | | | |
| Systolic BP, mmHg | 10 427 | 130.5 [0.12] | 125.2 [0.14] | −5.31 [0.17]*** | 124.8 [0.16] | −5.67 [0.18]*** |
| Diastolic BP, mmHg | 10 427 | 84.0 [0.07] | 79.6 [0.10] | −4.35 [0.11]*** | 79.5 [0.11] | −4.48 [0.12]*** |
| Weight, kg | 10 428 | 101.2 [0.09] | 99.0 [0.11] | −2.20 [0.06]*** | 97.6 [0.12] | −3.61 [0.08]*** |
| **Diabetes cohort** | | | | | | |
| HbA1c, % | 1738 | 8.06 [0.04] | 6.88 [0.03] | −1.18 [0.05]*** | 6.68 [0.03] | −1.38 [0.05]*** |
| Weight, kg | 1738 | 106.3 [0.24] | 102.9 [0.28] | −3.34 [0.17]*** | 101.0 [0.31] | −5.24 [0.22]*** |

**Abbreviations:** BP, blood pressure; EMM, estimated marginal mean; HbA1c, glycated hemoglobin; HDL, high-density lipoprotein; LDL, low-density lipoprotein; SE, standard error.

^a^ Change from baseline. *** p < 0.001; ** p < 0.01; * p < 0.05.

^b^ Models restricted to patients with available pharmacy data (~76–78% of each cohort). Dyslipidemia models (LDL, TC, HDL, weight) adjusted for statin and GLP-1 agonist use at enrollment; triglycerides model additionally adjusted for fibrate use at enrollment. Hypertension models adjusted for antihypertensive and GLP-1 agonist use at enrollment. Diabetes models adjusted for GLP-1 agonist, metformin, and insulin use at enrollment. All models additionally adjusted for age, sex, and BMI at enrollment.

**eTable 2.** Sensitivity analysis: primary models with race and ethnicity as covariate, patients with nonmissing race and ethnicity data^b^

| **Outcome** | **N** | **Baseline EMM [SE]** | **3 mo** | | **6 mo** | |
| --- | --- | --- | --- | --- | --- | --- |
|  |  |  | **EMM [SE]** | **Change [SE]^a^** | **EMM [SE]** | **Change [SE]^a^** |
| **Dyslipidemia cohort** | | | | | | |
| LDL cholesterol, mg/dL | 3112 | 134.9 [2.87] | 123.9 [2.89] | −10.97 [0.74]*** | 122.1 [2.90] | −12.80 [0.82]*** |
| Weight, kg | 3128 | 92.2 [0.73] | 89.8 [0.74] | −2.34 [0.10]*** | 88.5 [0.75] | −3.68 [0.13]*** |
| **Hypertension cohort** | | | | | | |
| Systolic BP, mmHg | 5674 | 131.3 [0.65] | 126.6 [0.65] | −4.75 [0.23]*** | 126.0 [0.66] | −5.32 [0.26]*** |
| Weight, kg | 5674 | 99.1 [0.57] | 96.8 [0.58] | −2.27 [0.08]*** | 95.5 [0.58] | −3.59 [0.11]*** |
| **Diabetes cohort** | | | | | | |
| HbA1c, % | 990 | 7.94 [0.12] | 6.78 [0.11] | −1.16 [0.06]*** | 6.60 [0.11] | −1.34 [0.07]*** |
| Weight, kg | 990 | 102.1 [0.96] | 99.0 [0.99] | −3.11 [0.22]*** | 96.8 [1.00] | −5.26 [0.30]*** |

**Abbreviations:** BP, blood pressure; EMM, estimated marginal mean; HbA1c, glycated hemoglobin; LDL, low-density lipoprotein; SE, standard error.

^a^ Change from baseline. *** p < 0.001; ** p < 0.01; * p < 0.05.

^b^ Models restricted to patients with nonmissing race and ethnicity data (~41–49% of each cohort), with race and ethnicity added as a categorical covariate. Analyses conducted for the condition-specific primary outcome and weight in each cohort. Restriction to patients with nonmissing race data introduces potential selection bias; results interpreted cautiously. All models additionally adjusted for age, sex, and BMI at enrollment. Results were consistent with primary model estimates across all outcomes.

**eTable 3.** Sensitivity analysis: primary models in data-availability-only cohort^b^

| **Outcome** | **N** | **Baseline EMM [SE]** | **3 mo** | | **6 mo** | |
| --- | --- | --- | --- | --- | --- | --- |
|  |  |  | **EMM [SE]** | **Change [SE]^a^** | **EMM [SE]** | **Change [SE]^a^** |
| **Dyslipidemia cohort** | | | | | | |
| LDL cholesterol, mg/dL | 11 296 | 135.7 [0.35] | 121.8 [0.39] | −13.89 [0.41]*** | 120.9 [0.45] | −14.78 [0.47]*** |
| Total cholesterol, mg/dL | 11 416 | 218.9 [0.36] | 199.3 [0.44] | −19.59 [0.45]*** | 199.0 [0.50] | −19.83 [0.51]*** |
| HDL cholesterol, mg/dL | 11 333 | 53.6 [0.14] | 52.4 [0.14] | −1.25 [0.11]*** | 53.0 [0.15] | −0.63 [0.12]*** |
| Triglycerides, mg/dL | 11 329 | 163.2 [0.77] | 136.4 [0.73] | −26.81 [0.76]*** | 135.8 [0.87] | −27.38 [0.88]*** |
| Weight, kg | 11 465 | 93.6 [0.08] | 91.4 [0.10] | −2.26 [0.06]*** | 90.2 [0.11] | −3.42 [0.08]*** |
| **Hypertension cohort** | | | | | | |
| Systolic BP, mmHg | 30 401 | 130.4 [0.07] | 125.7 [0.09] | −4.76 [0.10]*** | 125.5 [0.10] | −4.98 [0.11]*** |
| Diastolic BP, mmHg | 30 406 | 84.1 [0.04] | 79.9 [0.06] | −4.21 [0.07]*** | 79.7 [0.07] | −4.36 [0.07]*** |
| Weight, kg | 30 407 | 100.4 [0.05] | 98.7 [0.06] | −1.72 [0.04]*** | 97.7 [0.08] | −2.73 [0.06]*** |
| **Diabetes cohort** | | | | | | |
| HbA1c, % | 5101 | 8.13 [0.03] | 7.04 [0.02] | −1.09 [0.03]*** | 6.85 [0.02] | −1.28 [0.03]*** |
| Weight, kg | 5101 | 105.1 [0.14] | 102.2 [0.17] | −2.93 [0.11]*** | 100.5 [0.20] | −4.63 [0.15]*** |

**Abbreviations:** BP, blood pressure; EMM, estimated marginal mean; HbA1c, glycated hemoglobin; HDL, high-density lipoprotein; LDL, low-density lipoprotein; SE, standard error.

^a^ Change from baseline. *** p < 0.001; ** p < 0.01; * p < 0.05.

^b^ Models include all patients meeting condition-specific eligibility and cohort criteria with at least one follow-up outcome measurement, without requiring active engagement near 6 months. All models adjusted for age, sex, and BMI at enrollment. Results were consistent with or slightly larger than primary model estimates, confirming the engagement restriction did not inflate primary findings.

### **Figure Legends**

**eFigure 1. Three condition-specific cohorts, totaling 18,965 unique patients, were selected through sequential eligibility criteria.**

This flow diagram shows how the dyslipidemia, hypertension, and diabetes cohorts were selected from the full Nourish adult patient population with a first appointment before November 2025 (N = 319,497). The three cohorts are not mutually exclusive; patients meeting criteria for more than one condition appear in each applicable cohort. A medication sensitivity subset, limited to patients with complete pharmacy data, is also shown. Full definitions of eligibility, engagement, and outcome criteria are in the Methods section.

Abbreviation: RD, registered dietitian.
