## Supplementary figures and images for "Real-World Outcomes of Virtual Medical Nutrition Therapy in Dyslipidemia, Hypertension, or Diabetes"

### eFigure 1

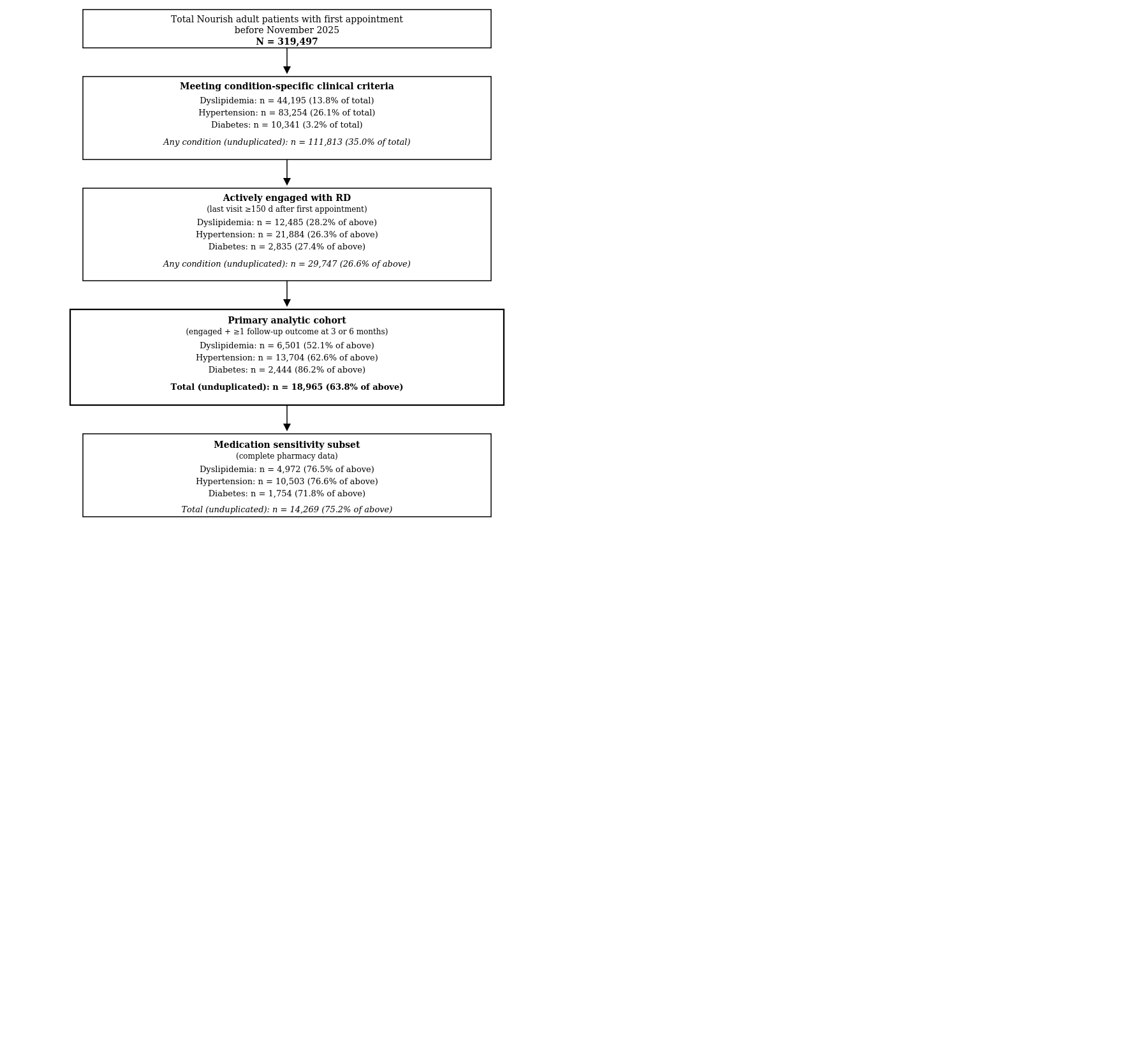
